# Validation of the Brief-Cope Questionnaire in a Seropositive Rheumatoid Arthritis Population

**DOI:** 10.64898/2026.08.28.26361589

**Authors:** Ioana Iliadis, Ivo Heitland, Kirsten Hoeper, Torsten Witte, Kai G. Kahl, Britta Stapel, Dirk Meyer-Olson

**Affiliations:** Department of Psychiatry, Social Psychiatry and Psychotherapy, Hannover Medical School, Hanover, Germany; Department of Rheumatology and Immunology, Hannover Medical School, Hanover, Germany

**Keywords:** rheumatoid arthritis, brief-cope, exploratory factor analysis

## Abstract

**Objective:** The Brief-cope questionnaire explore coping behavior. However, the underlying factor structure remains a subject of ongoing debate. Exploratory factor analyses (EFA) conducted across different populations have identified factor solutions ranging from two to fourteen factors. As of yet, the underlying factor structure of the Brief-cope has not been investigated in patients with seropositive rheumatoid arthritis (RA). Therefore, the aim of this study was to explore the underlying factor structure of the Brief-cope in a German population of seropositive RA.

**Methods:** 216 outpatients with seropositive RA completed the Brief-cope. An EFA with principal axis factoring and Promax rotation was conducted.

**Results:** EFA indicated a five-factor solution. The five-factor solution explained 51.95% of variance. The identified factors were: (1) problem-focused coping (Cronbach’s **α** = .851), (2) emotion-focused coping (**α** = .754), (3) maladaptive coping (**α** = .747), (4) religious coping (**α** = .851), and (5) substance-use coping (**α** = .869).

**Conclusion:** A five-factor solution provided the most appropriate representation of the underlying factor structure of the Brief-cope in patients with seropositive RA. This factor structure may serve as a suitable basis for future analyses of Brief-cope data in comparable RA populations.

**Trial registration number:** DRKS00013055.

## INTRODUCTION

The Brief-cope constitutes a commonly used instrument to assess coping behavior (1, 2). The questionnaire contains 28 items and is divided into 14 subscales (2). Numerous researchers have assessed the underlying factor structure of the Brief-cope (1, 3-7). Among the best-known solutions are the two-factor model by Eisenberg and colleagues who examined heart failure patients and the three-factor model by Dias and colleagues who analyzed athletes (6, 7). The model by Eisenberg and colleagues distinguishes between approach coping and avoidant coping (7). However, the subscales “humor” and “religion” could not be assigned to either factor (7). Another frequently applied model is the three-factor solution proposed by Dias and colleagues, comprising problem-focused coping, emotion-focused coping, and avoidant coping (6). Despite these proposed models, there is currently no consensus regarding the underlying factor structure of the Brief-cope (1, 3-5, 8). Several studies have demonstrated that the previously proposed factor solutions do not adequately fit their data and therefore recommend conducting an exploratory factor analysis (EFA) before further analyses are performed on respective samples (1-5, 8). Consequently, researchers across various disciplines frequently perform EFAs to identify the most appropriate factor structure for their respective study populations (3-5, 8). A meta-analysis by Solberg and colleagues, demonstrated the heterogeneity of factor analyses results, ranging between two to 14 factors, depending on study population (1). Thus, the factor structure of the Brief-cope remains a matter of ongoing debate. While some researchers continue to search for a global factor structure, others argue to carry out a factor analysis for each target population separately.

In rheumatology, coping has increasingly gained attention as an important factor associated with psychological outcomes and disease-related outcomes (9-11). Accordingly, various instruments, including the Brief-cope, have been used to assess different coping behaviors in patient with rheumatoid arthritis (RA) (9-11). However, the underlying factor structure of the German translation of the Brief-cope has not yet been investigated in a population of seropositive RA. Therefore, the aim of the present study was to explore the underlying factor structure of the Brief-cope in patients with seropositive RA.

## METHOD

### Study Design

The multicenter study was approved by the ethical committee of Hannover Medical School (number 3638– 2017) and was conducted in accordance to the ethical principles of the declaration of Helsinki (12). The study was carried out in eight rheumatology outpatient clinics in Germany, from 01/2018 to 12/2019. Participants were informed about the study, and all participants provided written informed consent before study inclusion. Participants received a 12 months long medical intervention program following a treat-to-target (T2T) approach, whereby participants had multiple follow-up visits (baseline and weeks 6 (week 8 optional), 12, 24, 36 and 52) with the rheumatologist. Thereby, Disease Activity Score in 28 Joints measured with C reactive protein (DAS28), relevant clinical data, as well as sociodemographic and socioeconomic were recorded at baseline and follow-up (13, 14).

### Subjects

In order to participate in the study, participants had to be at least 18 years old and need to be diagnosed with an anti-citrullinated protein antibodies positive result (ACPA/RF-positive) RA (American College of Rheumatology (ACR)/EULAR criteria 28) (13, 14). Participants were excluded from the study if they meet one of the following criteria: having severe comorbidities and insufficient German language skills.

### Outcome Measurements

The primary outcome measurement was the German version of the Brief-cope questionnaire (2). The questionnaire consists of 28 questions which are divided into 14 subscales: Active Coping (**α** = .68), Planning (**α** = .73), Positive Reframing (**α** = .64), Acceptance (**α** =.57), Humor (**α** =.73), Religion (**α** = .82), Using Emotional Support (**α** = .71), Using Instrumental Support (**α** = .64), Self-Distraction (**α** = .71), Venting (**α** = .50), Denial (**α** = .54), Substance Use (**α** = .90), Behavioral Disengagement (**α** = .65), and Self-Blame (**α** = .69) (2). Participants are asked to answer each item on a 4-point Likert-scale ranging from 1 (usually don’t do this at all) to 4 (I usually do this a lot).

### Statistical analysis

All statistical analyses were conducted using IBM SPSS Statistics Version 29. In order to examine the underlying factor structure, an EFA using principal axis factoring with oblique Promax rotation was conducted using baseline data, only. Promax rotation was used, because coping is expected to interrelate, and therefore independence of factors could not be assumed. In Addition, internal consistency was tested by calculating Cronbach’s alpha for all generated factors based on baseline data.

## RESULTS

### Study population

A total of 216 participants with seropositive RA were included in the analyses. Participants had a mean age of 58.56 years (*SD* = 11.8), and n = 161 (74.5%) were female. The mean DAS28 score was 4.57 (*SD* = 1.14). Detailed demographic and clinical characteristics are shown in Table 1.

**Table 1.**
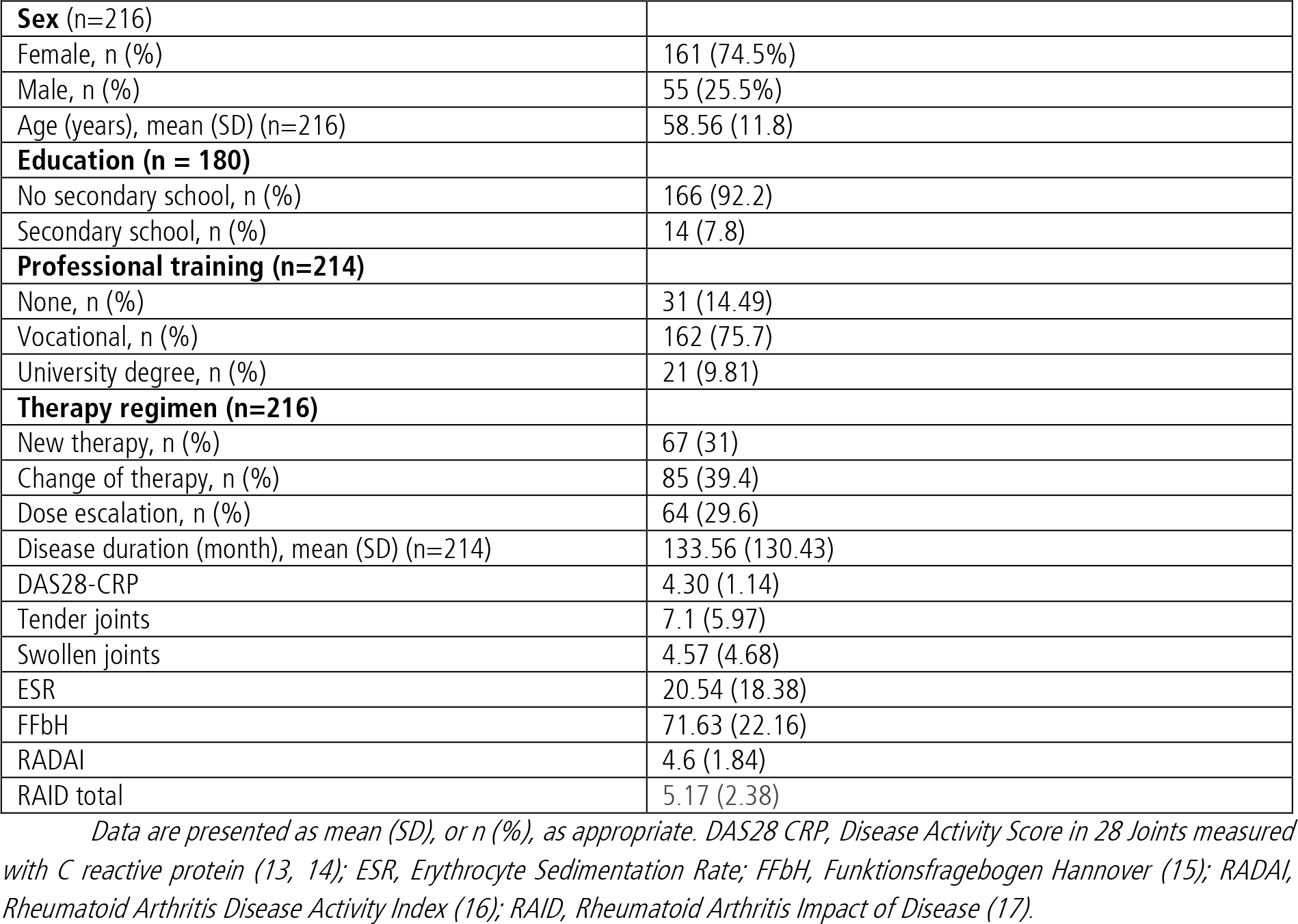
Baseline Characteristics of study population.

Data are presented as mean (SD), or n (%), as appropriate.
|  |  |
| --- | --- |
| **Sex (n=216)** |  |
| Female, n (%) | 161 (74.5%) |
| Male, n (%) | 55 (25.5%) |
| Age (years), mean (SD) (n=216) | 58.56 (11.8) |
| **Education (n = 180)** |  |
| No secondary school, n (%) | 166 (92.2) |
| Secondary school, n (%) | 14 (7.8) |
| **Professional training (n=214)** |  |
| None, n (%) | 31 (14.49) |
| Vocational, n (%) | 162 (75.7) |
| University degree, n (%) | 21 (9.81) |
| **Therapy regimen (n=216)** |  |
| New therapy, n (%) | 67 (31) |
| Change of therapy, n (%) | 85 (39.4) |
| Dose escalation, n (%) | 64 (29.6) |
| Disease duration (month), mean (SD) (n=214) | 133.56 (130.43) |
| DAS28-CRP | 4.30 (1.14) |
| Tender joints | 7.1 (5.97) |
| Swollen joints | 4.57 (4.68) |
| ESR | 20.54 (18.38) |
| FFbH | 71.63 (22.16) |
| RADAI | 4.6 (1.84) |
| RAID total | 5.17 (2.38) |

### Exploratory factor analysis

An EFA using principal axis factoring with Promax rotation was conducted to examine the underlying structure of the Brief-cope scale. The suitability of the data for EFA was confirmed by a Kaiser-Meyer-Olkin measure of sampling adequacy of .766 and a significant Bartlett’s test of sphericity (χ^2^(378) = 2280.11, *p* < .001), indicating that the correlation matrix was appropriate for factor analysis.

The Kaiser criterion suggested an eight-factor solution explaining 63.97% of the total variance. However, scree plot inspection supported a five-factor solution (see Figure 1). The five-factor model explained 51.95% of the total variance and comprised the following dimensions: Factor 1 (problem-focused coping) comprising 10 items and explaining 21.45% of the variance. Factor 2 (emotion-focused coping) comprising 7 items and further explaining 10.11% of the variance. Factor 3, maladaptive coping, comprising 5 items and explaining 7.97% of the variance. Factor 4, religious coping explaining 5.63% and Factor 5, substance-use coping, explaining 6.81% of the variance and comprising 2 items each. Items 6 and 16 were excluded as their factor loadings were below .30. Overall, factor loadings ranged from .42 to .88, and Cronbach’s alpha coefficients indicated acceptable to good internal consistency across all five factors (**α** = .75 - .87). Detailed factor loadings, explained variance, and reliability coefficients are presented in Table 2.

**Table 2.** Pattern matrix, explained variance and internal consistency of the five-factor solution.

Pattern matrix of the five-factor solution, explained variance per factor, and Cronbach's alpha coefficients per factor. Results were obtained from baseline-data only. Factors marked in bold indicates to which factor the item belongs.
| Brief-cope Items |  | Factor 1 | Factor 2 | Factor 3 | Factor 4 | Factor 5 |
| --- | --- | --- | --- | --- | --- | --- |
| 1 | I've been turning to work or other activities to take my mind off things. | -.037 | .183 | **.475** | .193 | .096 |
| 2 | I've been concentrating my efforts on doing something about the situation I'm in. | **.417** | .206 | .246 | -.081 | .032 |
| 3 | I've been saying to myself "this isn't real". | .067 | -.084 | **.632** | -.106 | -.140 |
| 4 | I've been using alcohol or other drugs to make myself feel better | -.007 | -.047 | -.006 | -.047 | **.881** |
| 5 | I've been getting emotional support from others. | **.580** | .001 | .065 | -.008 | -.085 |
| 6 | I've been giving up trying to deal with it. | -.187 | .269 | .165 | .005 | -.108 |
| 7 | I've been taking action to try to make the situation better. | **.446** | .297 | -.086 | .119 | -.038 |
| 8 | I've been refusing to believe that it has happened. | .099 | -.196 | **.728** | -.028 | -.104 |
| 9 | I've been saying things to let my unpleasant feelings escape. | **.474** | -.029 | .231 | -.121 | -.127 |
| 10 | I've been getting help and advice from other people. | **.825** | -.170 | -.098 | .028 | .008 |
| 11 | I've been using alcohol or other drugs to help me get through it. | -.006 | -.102 | -.013 | .010 | **.868** |
| 12 | I've been trying to see it in a different light, to make it seem more positive. | .146 | **.654** | -.169 | .015 | -.040 |
| 13 | I've been criticizing myself. | .027 | .070 | **.575** | .002 | .156 |
| 14 | I've been trying to come up with a strategy about what to do. | **.573** | .320 | -.005 | -.040 | .089 |
| 15 | I've been getting comfort and understanding from someone. | **.779** | -.066 | .033 | -.055 | -.089 |
| 16 | I've been giving up the attempt to cope. | -.229 | .074 | .155 | .015 | -.143 |
| 17 | I've been looking for something good in what is happening. | .063 | **.594** | -.065 | .068 | -.018 |
| 18 | I've been making jokes about it. | -.135 | **.461** | .218 | -.304 | .094 |
| 19 | I've been doing something to think about it less, such as going to movies, watching TV, reading, daydreaming, sleeping, or shopping. | .027 | **.513** | .202 | .095 | -.006 |
| 20 | I've been accepting the reality of the fact that it has happened. | -.056 | **.503** | .014 | .016 | -.029 |
| 21 | I've been expressing my negative feelings. | **.423** | -.179 | .221 | .037 | .080 |
| 22 | I've been trying to find comfort in my religion or spiritual beliefs. | -.030 | .003 | .064 | **.880** | -.043 |
| 23 | I've been trying to get advice or help from other people about what to do. | **.727** | -.070 | -.134 | .059 | .066 |
| 24 | I've been learning to live with it. | -.002 | **.629** | -.111 | .040 | -.147 |
| 25 | I've been thinking hard about what steps to take. | **.371** | .172 | .075 | .044 | .166 |
| 26 | I've been blaming myself for things that happened. | -.137 | -.036 | **.652** | .166 | .046 |
| 27 | I've been praying or meditating. | .009 | .014 | .043 | **.784** | .009 |
| 28 | I've been making fun of the situation. | -.177 | **.641** | -.124 | -.101 | .035 |
| Variance, % |  | 21.45 | 10.11 | 7.97 | 5.63 | 6.81 |
| Cronbach's Alpha at Baseline |  | .851 | .754 | .747 | .851 | .869 |

**Figure 1.**
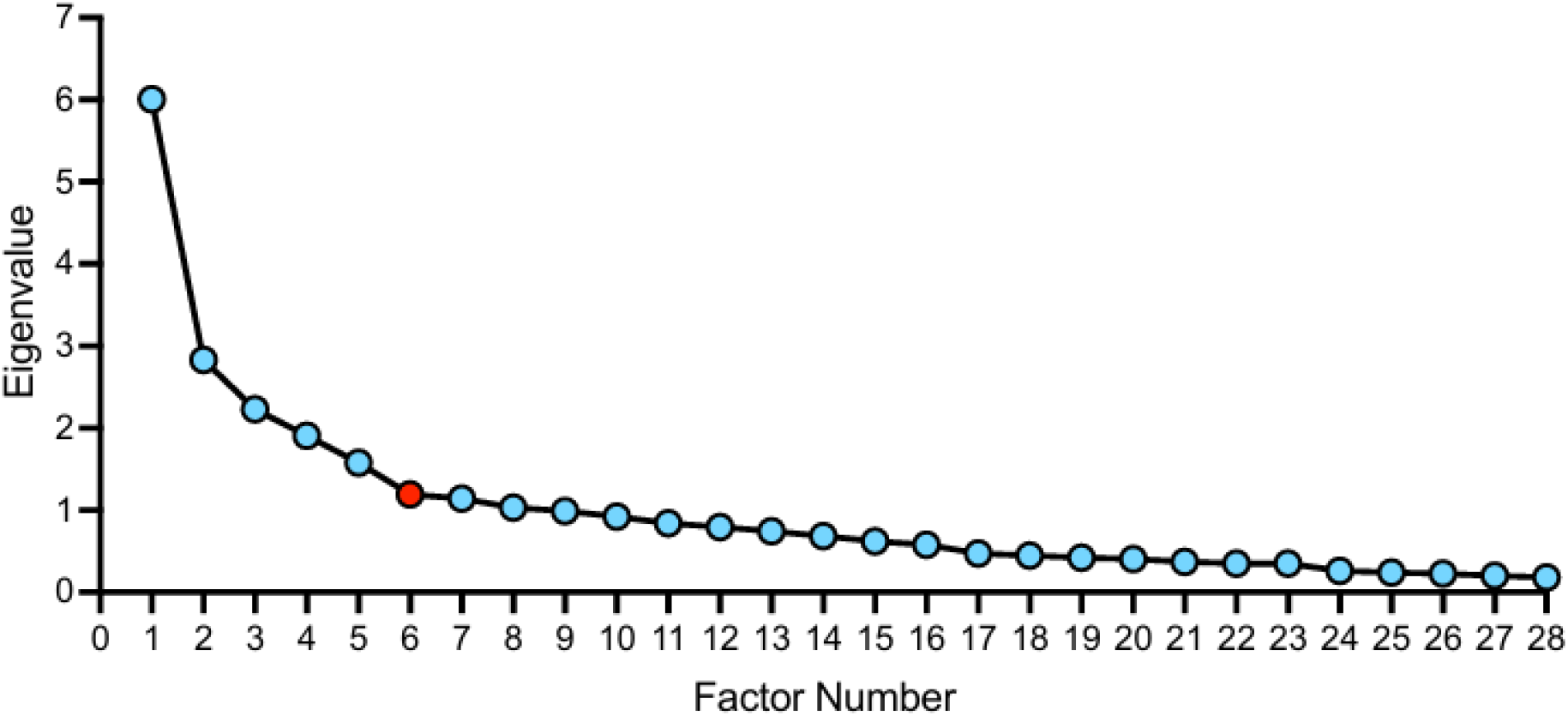
Screeplot Exploratory Factor Analysis Scree plot of the exploratory factor analysis (principal axis factoring) of the Brief-cope. The elbow of the curve supported retention of five factors.

## DISCUSSION

Results of the EFA indicated an eight-factor solution according to the Kaiser criterion, whereas scree plot inspection supported a five-factor solution. Since the Kaiser criterion is known to frequently overestimate the number of factors, it is generally considered less reliable for determining the optimal factor solution (18). In contrast, the five-factor solution provided a more parsimonious and conceptually coherent representation of coping strategies with acceptable to good internal consistency across all identified factors. Thus, the first five factors explained 51.95% of the total variance, whereas the additional three factors increased the explained variance by only 12.01%. Consequently, it is recommended to use the five-factor solution in further analyses for RA populations.

To date, no universally accepted factor structure for the Brief-cope has been established (1, 3-5, 19, 20). According to a recent meta-analysis, factor structure of the Brief-cope ranges from two to fourteen factors with a mean of 5.33 factors (1). Thereby, it is important to acknowledge that all studies were conducted in different populations and across different cultural and linguistic settings. These differences may contribute to the observed heterogeneity in Brief-cope factor structures. Consequently, our findings highlight the heterogeneity of factor solution as well and support the recommendation to evaluated factor structure within the studied population rather than assuming the universal applicability of an existing model. To our knowledge, this is the first study to investigate the underlying factor structure of the Brief-cope in a German population of patients with seropositive RA. Although Wrobel and colleagues applied the Brief-cope in patients with RA, they did not perform a factor analysis before conducting subsequent analyses (11).

This study has some limitations. First, the proposed factor structure was derived from EFA and was not confirmed using confirmatory factor analysis (CFA). Therefore, future studies should apply a CFA to validate the identified five-factor solution in an independent sample. Second, the present findings are based on a German cohort of patients with seropositive RA and may therefore not be generalized to other patient populations or cultural or linguistic settings.

## CONCLUSION

To our best knowledge, this is the first study testing the underlying factor structure of the Brief-cope in a German, seropositive RA population. Study results revealed a five-factor solution for this population. While the five-factor model appears to be applicable for seropositive RA patients, results also support the recommendation that the factor structure of the Brief-cope should be evaluated within the given studied population priorly rather than assuming the universal applicability of an existing model.

## Data Availability

All data produced in the present study are available upon reasonable request to the authors.

## Acknowledgements

We would like to thank all participating patients for their willingness to contribute to the study.

